# Reference values for amplitude-integrated EEGs in children from 1 month to 17 years of age

**DOI:** 10.1101/2022.03.18.22272592

**Authors:** Sandra Greve, Verena Tamara Löffelhardt, Adela Della-Marina, Ursula Felderhoff-Müser, Christian Dohna-Schwake, Nora Bruns

## Abstract

**Aim:** Amplitude-integrated electroencephalography (aEEG) is used to monitor electrocortical activity in critically ill children, but reference values are lacking for patients older than 3.5 months. We aimed to derive reference values for paediatric aEEGs from neurologically healthy children.

**Methods:** Normal EEGs from awake children aged 1 month to 17 years (213 female, 237 male) without neurological disease or neuroactive medication were retrospectively converted into aEEGs. Two observers manually measured the upper and lower amplitude borders of the C3 – P3, C4 – P4, C3 – C4, P3 – P4, and Fp1 – Fp2 channels of the 10 – 20 system. Percentiles (10^th^, 25^th^, 50^th^, 75^th^, 90^th^) were calculated for each age group (< 1 year, 1 year, 2 – 5 years, 6 – 9 years, 10 – 13 years, 14 – 17 years).

**Results:** Amplitude heights and curves differed between channels without sex-specific differences. During the first 2 years of life, upper and lower amplitudes of all but the Fp1 – Fp2 channel increased and then declined until 17 years. The decline of the upper Fp1 – Fp2 amplitude began at four years, while the lower amplitude declined from the first year of life.

**Interpretation:** aEEG interpretation must account for age and electrode positions but not for sex in infants and children.

**What this paper adds:**

- aEEG amplitudes increase in the first years of life and then decline.
- aEEG amplitudes depend on electrode positions but not on sex.
- aEEG interpretation must account for age and electrode positions.

---

In recent years, the use of amplitude-integrated EEG (aEEG) has expanded from neonatology into paediatric intensive care, driven by the need for continuous neurophysiological monitoring and the advantages of aEEG as an affordable, broadly available, and easy-to-interpret bedside technique (2). aEEG has proven useful for the assessment of seizures and guiding antiepileptic treatment in critically ill children (3-7). There is incipient but growing evidence that physiological and pathological conditions induce changes in the background pattern, e.g., sleep states, sedation, cardiac arrest, central nervous system infections, and inflammation (4, 5, 8-13). Normalization of background patterns according to a neonatal classification (14) has been described to be predictive of outcome in neonates, children, and adults after hypoxic events (9, 15-17).

Neonatal aEEG classifications found that the physiological aEEG background rises with gestational and postnatal age (18-21). Despite these findings and increasing aEEG use in older infants and children, reference values have not been defined for infants and children above 3.5 months of age. Specific diseases and interventions in paediatric intensive care can require adaptations to the standard electrode positions used in neonatology, making the establishment of reference values more complex than in neonates.

To address the growing need for reference values (22), we calculated aEEGs from normal EEGs recorded in awake children without cerebral disease who did not receive sedatives, antiepileptic drugs or any other type of neuroactive medication. The aim was to provide reference values for bedside assessment of aEEG in the PICU. We measured the upper and lower margins of five aEEG channels that are used in paediatric intensive care and calculated age-specific percentiles.

## Methods

EEGs without pathologies from awake children between 1 month and 17.9 years of age and without central nervous system disease or neuroactive medication were eligible for the study. EEGs were classified into one-year age groups during the selection process. To avoid bias from repeated recordings in the same patient, only one EEG per one-year age group was selected from each patient.

### Ethics approval

The study was approved by the local ethics committee of the Medical Faculty of the University of Duisburg-Essen (20-9444-BO). Informed consent was not necessary according to local legislation because retrospective anonymized data were used.

### Indication for EEG recording and verification of eligibility

Most EEGs were conducted as part of the clinical routine in several conditions before the initiation of therapy (Table 1). Our centre has a large paediatric oncology department. All children undergoing chemotherapy receive an EEG before therapy in order to have a baseline finding in case of neurological complications. Another specialty of our centre are solid organ transplants and bone marrow transplants (oncological and non-oncological). The same pre-therapy diagnostics are applied to these patients. Some other indications were diagnostic work- ups in suspected neurologic disease. No inborn neurologic disease was allowed to be diagnosed at any timepoint in the patient history. For acquired brain injury, only recordings before the insult were eligible.

**Table 1:** Eligibility criteria

| Inclusion criteria (all must be met) | Exclusion criteria (one is sufficient) |
| --- | --- |
| • Age < 18 years<br>• EEG normal according routine assessment<br>• Awake throughout recording | • Acquired or inborn cerebral disease or damage<br>• Antiepileptic drug use or other neuroactive medication<br>• Former preterm infant before 24 months corrected age<br>• Fallen asleep during recording<br>• Developmental delay<br>• Photostimulation during recording* |
| In case of more than one recording within the same year of age: only one random EEG used to avoid bias from repeated measurements in the same subject |  |

\*Hyperventilation was not an exclusion criterion because all EEGs were annotated. Amplitudes were only assessed until the beginning of hyperventilation.

Patient histories were checked for neuroactive substances and patients excluded if they received any type of neuroactive medication at the time of recording.

### EEG recording

Full-channel EEG was applied according to the international 10-20 system after skin preparation with OneStep EEG Gel Abrasiv plus®. An impedance check was performed, and skin preparation was repeated until impedances < 50 kΩ were achieved for all electrodes, according to requirements for standard EEG recording.

### EEG interpretation

The EEG reader was a board-certified paediatric neurologist (ADM) with additional certificates in EEG and epileptology by the German Society for Epileptology (DGfE). Because all EEGs were interpreted by ADM or, on rare occasions, by a substitute who holds the same qualifications, no additional independent read was performed before inclusion of EEGs into the study.

### EEG conversion

aEEGs were calculated using Polaris EEG software (Polaris Trend Software QP-160AK, Nihon Kohden, Tokyo, Japan). To obtain the aEEG, the raw EEG signal was amplified, passed through an asymmetric bandpass filter, logarithmically transformed, and rectified as described by Maynard et al. (23).

Channels C3 – C4, P3 – P4, C3 – P3, C4 – P4, and Fp1 – Fp2 of the 10 – 20 system were converted (Figure 1a). All channels except for the Fp1-Fp2 channel are standard for aEEG conduction in neonates. The Fp1 – Fp2 channel was additionally included because in patients with head injury or after neurosurgery, the frontal positions can be the only available locations for electrode placement.

**Figure 1:**
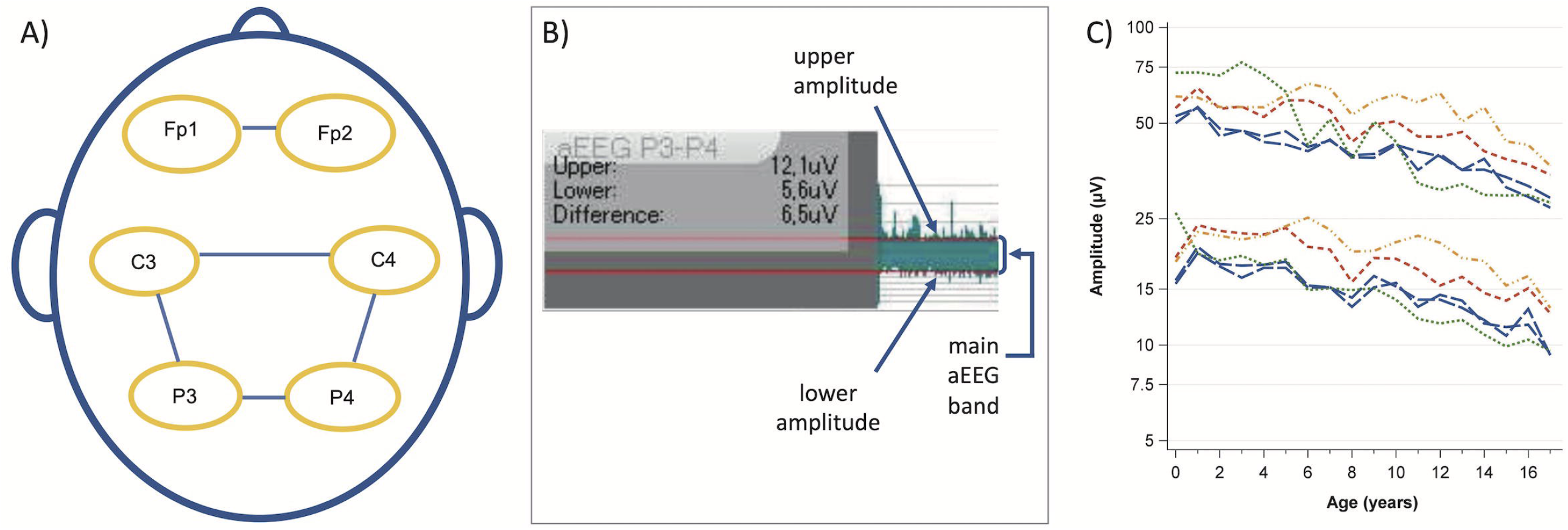
A) Analysed channels according to the 10 – 20 system. B) Amplitude assessment using the integrated ruler for amplitude measurement. The red lines were manually aligned with the main aEEG band. The small spikes above and below the main band were not considered. C) Evolution of the median upper and lower amplitude values with age. Blue long-dashed line: C3 – P3 and C4 – P4, red medium dashed line: C3 – C4, green short-dashed line: Fp1 – Fp2, orange dashed-dotted line: P3 – P4.

### aEEG analysis

SG and NB measured the upper and lower amplitude borders manually with the integrated tool for amplitude measurement as previously described (13) (Figure 1b).

An image of the measured tracing was saved as a PDF file for documentation purposes and quality control. The measured values were transferred manually into an Excel spreadsheet.

### Data analysis

Categorical variables are summarized as counts and relative frequencies. aEEG amplitudes are presented as age-specific percentiles (10^th,^ 25^th^, 50^th^, 75^th^, and 90^th^). Other continuous variables are presented as means and 95% confidence intervals (CI) or standard deviation (SD) if normally distributed and as median and interquartile range (IQR) if non-normally distributed.

#### Calculation of amplitudes

For each aEEG channel, we calculated the mean of the values measured by the two raters for the upper and lower amplitudes.

#### Definition of age groups

The raw data were visualized to assess the height and evolution of amplitudes with age (Figure 1c). Next, we defined age groups to calculate percentiles (< 1 year, 1 year, 2 – 5 years, 6 – 9 years, 10 – 13 years, and 14 – 17 years). The aim was to depict the rapid amplitude changes during the first years of life, provide sufficient detail on amplitude differences at older ages, and avoid an excessive number of subgroups. The C3 – P3 and C4 – P4 channels were collapsed for percentile calculations because they represent the contralateral positions during 2-channel recordings. To assess sex-specific differences, we calculated means and 95 % CIs for males and females for each channel and age group.

#### Interrater reliability

Bland–Altman plots (24) were created for the upper and lower amplitudes of each channel for each year of age to rule out systematic differences in measurements between the two raters. Mean differences and SD of the differences between the two raters were calculated for the upper and lower borders. Intraclass correlation coefficients for the upper and lower borders were calculated using a two-way mixed model for individual ratings (ICC 3,1) (25).

#### Software

SAS Enterprise Guide 8.4 (SAS Institute Inc., Cary, NC, USA) was used to perform statistical analyses and produce figures.

## Results

Out of 11,543 EEGs recorded between January 2014 and February 2020 in the Children’s Hospital of the University Hospital Essen, 450 unremarkable EEGs were included (Figure 2). The median (IQR) duration was 17 (15 - 18) minutes. Males accounted for 52.7 % of the measurements and 60.7 % of EEGs were conducted as routine diagnostics (Table 2).

**Table 2:** Included patients and indications for EEG

|  | **n (%)** | **Mean (SD)** |
| --- | --- | --- |
| Male | 237 (52.7) |  |
| Female | 213 (47.3) |  |
| Age group |  |  |
| < 1 year | 44 (9.8) | 6 months (4 months) |
| 1 year | 30 (6.7) | 1 year 6 months (4 months) |
| 2-5 years | 96 (21.3) | 3 years 7 months (14 months) |
| 6-9 years | 79 (17.6) | 7 years 10 months (14 months) |
| 10-13 years | 86 (19.1) | 12 years 2 months (14 months) |
| 14-17 years | 115 (25.6) | 16 years 1 month (14 months) |
| Indications |  |  |
| Routine diagnostics* | 273 (60.7) |  |
| Diagnostic work-up in patients with suspected inborn or acquired neurologic disease** | 177 (39.3) |  |

\*before the initiation of therapy or during aftercare, e.g. before or after solid organ transplant, bone marrow transplant, or chemotherapy
\*\* no neurologic disease diagnosed after completion of diagnostics at any timepoint in the patient history (inborn) or before the recording (acquired)

**Figure 2:**
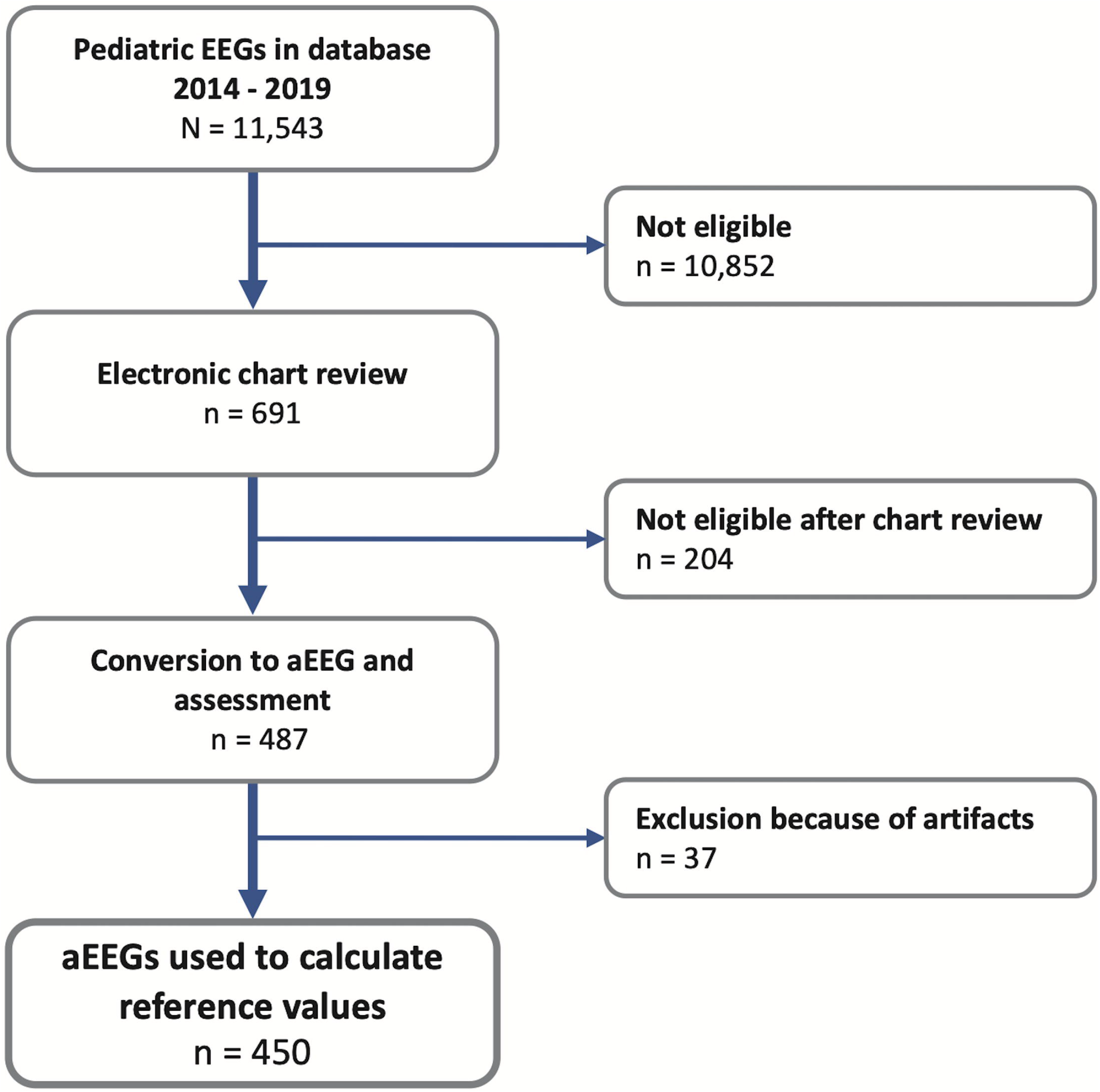
Flow chart of EEG selection and aEEG conversion.

The intraclass correlation coefficient was 0.83 for the upper and 0.87 for the lower amplitude. The mean interrater difference for the upper amplitude was 0.9 µV (SD 6.3 µV) and 0.3 µV (2.2 µV) for the lower amplitude.

In the C3 – P3 and C4 – P4 channels, the amplitudes rose from 0 to 1 year of age and showed a continuous decline until the age of 17 years (Figure 3a). A similar trend was observed in all channels, but there were differences in amplitude values and curves between the channels (Table 3, figure 3 b-d). We found statistically non-significant differences in mean amplitudes between males and females (Supplementary table).

**Table 3:** Percentiles by channels and age groups

| Channel | Age | Amplitude | Percentile [μV] | | | | |
| --- | --- | --- | --- | --- | --- | --- | --- |
|  |  |  | 10th | 25th | 50th | 75th | 90th |
| C3-P3/<br>C4-P4 | < 1 year | Lower | 11 | 13 | **16** | 21 | 25 |
|  |  | Upper | 34 | 42 | **51** | 62 | 97 |
|  | 1 year | Lower | 15 | 18 | **20** | 23 | 25 |
|  |  | Upper | 40 | 48 | **56** | 62 | 77 |
|  | 2 - 5 years | Lower | 12 | 15 | **18** | 20 | 25 |
|  |  | Upper | 30 | 39 | **46** | 56 | 67 |
|  | 6 - 9 years | Lower | 11 | 12 | **15** | 19 | 24 |
|  |  | Upper | 28 | 35 | **42** | 53 | 60 |
|  | 10 - 13 years | Lower | 9 | 10 | **14** | 19 | 23 |
|  |  | Upper | 24 | 29 | **38** | 52 | 61 |
|  | 14 - 17 years | Lower | 7 | 9 | **11** | 15 | 19 |
|  |  | Upper | 17 | 23 | **31** | 40 | 52 |
| C3-C4 | < 1 year | Lower | 14 | 16 | **19** | 24 | 27 |
|  |  | Upper | 43 | 50 | **56** | 68 | 82 |
|  | 1 year | Lower | 20 | 22 | **24** | 27 | 28 |
|  |  | Upper | 51 | 55 | **65** | 77 | 85 |
|  | 2 - 5 years | Lower | 16 | 19 | **23** | 26 | 31 |
|  |  | Upper | 41 | 47 | **56** | 70 | 81 |
|  | 6 - 9 years | Lower | 14 | 15 | **19** | 23 | 28 |
|  |  | Upper | 34 | 41 | **53** | 63 | 72 |
|  | 10 - 13 years | Lower | 12 | 14 | **17** | 22 | 26 |
|  |  | Upper | 30 | 37 | **49** | 57 | 74 |
|  | 14 - 17 years | Lower | 9 | 11 | **14** | 17 | 21 |
|  |  | Upper | 23 | 29 | **37** | 49 | 58 |
| P3-P4 | < 1 year | Lower | 13 | 14 | **18** | 22 | 27 |
|  |  | Upper | 39 | 47 | **61** | 72 | 106 |
|  | 1 year | Lower | 17 | 19 | **23** | 25 | 29 |
|  |  | Upper | 48 | 53 | **60** | 77 | 85 |
|  | 2 - 5 years | Lower | 16 | 19 | **22** | 26 | 32 |
|  |  | Upper | 42 | 49 | **57** | 70 | 86 |
|  | 6 - 9 years | Lower | 15 | 17 | **21** | 27 | 33 |
|  |  | Upper | 39 | 48 | **61** | 74 | 93 |
|  | 10 - 13 years | Lower | 14 | 16 | **21** | 26 | 31 |
|  |  | Upper | 35 | 43 | **53** | 73 | 90 |
|  | 14 - 17 years | Lower | 9 | 12 | **15** | 20 | 26 |
|  |  | Upper | 26 | 31 | **43** | 59 | 71 |
| Fp1-Fp2 | < 1 year | Lower | 11 | 14 | **16** | 23 | 25 |
|  |  | Upper | 36 | 59 | **72** | 96 | 106 |
|  | 1 year | Lower | 14 | 16 | **19** | 23 | 27 |
|  |  | Upper | 49 | 62 | **72** | 86 | 129 |
|  | 2 - 5 years | Lower | 14 | 16 | **19** | 23 | 25 |
|  |  | Upper | 47 | 55 | **71** | 88 | 113 |
|  | 6 - 9 years | Lower | 11 | 13 | **15** | 16 | 20 |
|  |  | Upper | 33 | 38 | **44** | 56 | 75 |
|  | 10 - 13 years | Lower | 9 | 10 | **12** | 14 | 17 |
|  |  | Upper | 22 | 28 | **34** | 45 | 53 |
|  | 14 - 17 years | Lower | 8 | 9 | **10** | 12 | 14 |
|  |  | Upper | 21 | 24 | **29** | 37 | 45 |

**Figure 3:**
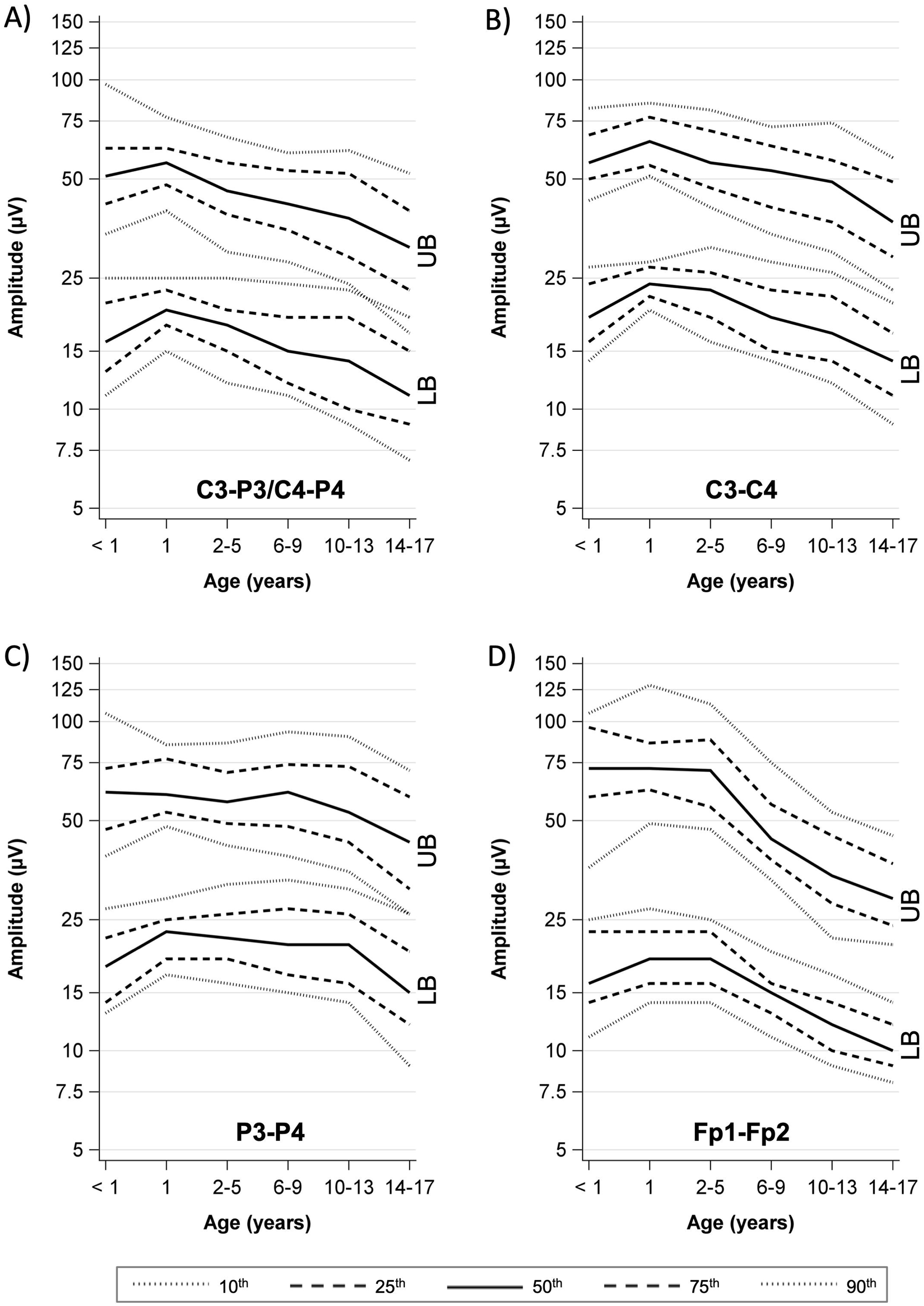
Percentile curves by age. A) C3-P3/C4-P4 channel. B) C3-C4 channel. C) P3-P4 channel. D) Fp1-Fp2 channel.

To facilitate bedside interpretation of aEEG, we created a pocket card with a map of the channels according to the 10 – 20 system, graphical percentiles for each channel, and summary tables with numerical percentile values (Supplementary file).

## Discussion

This is the first study to derive age-specific aEEG reference values in awake children without underlying or acquired cerebral disease. aEEG amplitudes increased during the first years of life, followed by a continuous decline until the age of 17 years. Different amplitude values and curves, but no sex-specific differences were observed between the channels.

The lower margin at all ages and for all channels had a 10^th^ percentile above 5 µV, corresponding to a continuous normal voltage background according to Hellström-Westas et al. (14). This neonatal classification is used up to adult age due to the lack of age-specific reference values (5, 7, 9, 17, 22, 26, 27). According to our data, a normal paediatric aEEG amplitude from any of the analysed channels fulfils the continuous normal voltage criteria by Hellström-Westas. Therefore, defining “normal” based on this classification in the paediatric aEEG studies mentioned above seems justified. The different amplitudes we found between channels show the impact of electrode positions on the aEEG background. This should be considered when comparing study findings and designing future investigations.

The effect of pathologies and sedatives on the aEEG background remains largely uninvestigated. In conventional EEG, abnormal oscillatory patterns are induced by anaesthetic and sedative drugs (28). These patterns reflect a patient’s current level of anaesthesia, making it possible to track the brain states of a patient (28). Intracranial injuries, a missing skullcap after decompressive craniectomy, modified electrode positions due to head dressings, and external injuries to the head can impact the raw EEG pattern and consecutively the aEEG amplitudes obtained. This calls for caution when applying aEEG reference values in patients with intracranial injury. However, in the case of side differences in aEEG channels, the percentiles may help to distinguish the normal side from the altered side, e.g., in patients with intracranial haemorrhage. With respect to background pattern changes over time, the percentiles may help to recognize deterioration or normalization.

The main limitation of our study is the fact that the reference values were derived from awake children. Many children undergoing aEEG monitoring in the PICU receive sedatives, suffer from intracranial pathologies or fall asleep during part of the recording. We previously showed that amplitudes differ between deep and light sleep in healthy children (13, 29), making the establishment of reference values during sleep substantially more difficult. For this reason, we focused on awake aEEGs as a first step toward a systematic aEEG assessment. Intracranial injuries such as large subdural hematoma or trapped air after neurosurgery make the expected aEEG amplitudes unpredictable. This must be considered when using the reference values for interpretation, as well as the influence of sedatives and other neuroactive medication. aEEG conversion algorithms differ between manufacturers and are not openly accessible. Therefore, the same aEEG tracing may have slightly different amplitudes depending on the software used. Further, the manual amplitude measurement may yield different results compared to an automated electronic assessment, limiting the comparability of results between assessment techniques. A disadvantage of automated assessment is the poor performance of software in artifact recognition and removal (30). In contrast, intensive care providers can easily ignore sections containing artifacts during visual bedside assessment.

The findings of this study point out the need to account for age and electrode positions when interpreting aEEGs in infants and children. Further research must determine if amplitude deviations from the here-defined percentiles are associated with pathologies, medication, cerebral dysfunction or outcomes.

## Conclusion

This study provides aEEG reference values for bedside assessment derived from awake children without cerebral pathology. The percentiles provided can contribute to the identification of normal and patterns and deviations. Patient age and electrode position, along with medications and pathologies that may affect electrocortical activity, must be considered upon interpretation.

## Supporting information

STROBE checklist

Pocket card

## Data Availability

All data produced in the present study are available upon reasonable request to the authors

## Acknowledgments

We would like to thank the EEG technical assistants of the Department of Paediatric Neurology of the University Hospital Essen for their support.

The study received funding from the Medical Faculty of the University of Duisburg-Essen (Corona Care program) and from the Stiftung Universitätsmedizin Essen. NB received funding from the Medical Faculty of the University of Duisburg-Essen (IFORES program) and from the Stiftung Universitätsmedizin Essen. The funder did not participate in the work.

The authors have no conflicts of interest relevant to this article to disclose.

## Authors’ contributions

NB conceptualized the study, acquired funding, collected and analyzed the data, drafted the initial manuscript, and revised the manuscript. SG and VL collected and interpreted data, participated in the drafting of the initial manuscript. ADM interpreted the original EEG. CDS and UFM, helped to conceptualize analyses and interpret data. All authors critically revised the manuscript for important intellectual content, approved the final manuscript as submitted, and agree to be accountable for all aspects of the work.

---

**Supplementary table:** Mean (95 % CI) amplitude values (µV) by sex and age group

| Agegroup | Sex | n | Channel<br>C – P |  | P3 – P4 |  | C3 – C4 |  | Fp1 – Fp2 |  |
| --- | --- | --- | --- | --- | --- | --- | --- | --- | --- | --- |
|  |  |  | Lower | Upper | Lower | Upper | Lower | Upper | Lower | Upper |
| < 1 year | Male | 24 | 16.0<br>(14.6 – 17.3) | 54.7<br>(47.8 – 61.5) | 18.5<br>(16.3 – 20.7) | 65.4<br>(54.6 – 76.3) | 18.9<br>(17.2 – 20.7) | 60.9<br>(51.4 – 70.4) | 18.0<br>(15.4 – 20.6) | 82.6<br>(71.2 – 94.1) |
|  | Female | 20 | 19.2<br>(16.7 – 21.7) | 60.5<br>(51.6 – 69.4) | 20.1<br>(17.0 – 23.2) | 63.1<br>(51.4 – 74.9) | 20.7<br>(17.9 – 23.4) | 63.2<br>(52.8 – 73.5) | 17.2<br>(14.6 – 19.8) | 66.3<br>(55.6 – 77.0) |
| 1 year | Male | 21 | 20.2<br>(19.0 – 21.3) | 55.4<br>(51.8 – 59.0) | 22.5<br>(20.5 – 24.4) | 60.8<br>(55.5 – 66.2) | 24.3<br>(22.8 – 25.8) | 65.2<br>(59.5 – 70.8) | 19.3<br>(17.2 – 21.4) | 82.0<br>(67.0 – 97.1) |
|  | Female | 9 | 21.2<br>(18.5 – 23.8) | 60.4<br>(51.2 – 69.6) | 23.5<br>(19.3 – 27.6) | 71.4<br>(56.3 – 86.6) | 24.7<br>(19.3 – 30.0) | 72.0<br>(50.9 – 93.0) | 19.8<br>(16.0 – 23.5) | 73.2<br>(62.0 – 84.4) |
| 2 – 5 years | Male | 46 | 18.4<br>(17.3 – 19.6) | 49.1<br>(45.8 – 52.5) | 22.9<br>(20.6 – 25.2) | 60.6<br>(54.2 – 67.0) | 23.7<br>(21.7 – 25.7) | 61.5<br>(54.8 – 68.1) | 19.4<br>(18.0 – 20.9) | 74.4<br>(67.5 – 81.4) |
|  | Female | 50 | 17.8<br>(17.0 – 18.7) | 47.0<br>(44.7 – 49.3) | 23.1<br>(21.6 – 24.6) | 60.5<br>(56.3 – 64.6) | 22.3<br>(21.0 – 23.5) | 57.8<br>(54.0 – 61.6) | 19.4<br>(17.7 – 20.4) | 75.9<br>(68.1 – 83.7) |
| 6 – 9 years | Male | 49 | 16.7<br>(15.7 – 17.6) | 45.8<br>(43.0 – 48.6) | 23.7<br>(21.9 – 25.6) | 65.7<br>(60.2 – 71.2) | 20.8<br>(19.2 – 22.4) | 55.8<br>(51.6 – 60.1) | 15.4<br>(14.5 – 16.3) | 50.7<br>(46.3 – 55.1) |
|  | Female | 30 | 15.3<br>(14.0 – 16.7) | 41.7<br>(38.4 – 44.9) | 21.1<br>(18.6 – 23.7) | 58.7<br>(51.9 – 65.5) | 18.6<br>(16.6 – 20.6) | 50.5<br>(45.0 – 56.1) | 14.1<br>(12.9 – 15.2) | 44.5<br>(39.0 – 50.0) |
| 10 – 13 years | Male | 46 | 16.0<br>(14.5 – 17.5) | 42.0<br>(38.4 – 45.5) | 23.0<br>(20.2 – 25.8) | 61.2<br>(54.0 – 68.3) | 18.8<br>(17.1 – 20.6) | 50.8<br>(45.7 – 55.9) | 12.8<br>(11.8 – 13.7) | 38.8<br>(34.7 – 43.0) |
|  | Female | 40 | 14.3<br>(13.1 – 15.4) | 39.1<br>(36.0 – 42.3) | 21.4<br>(18.8 – 24.0) | 57.6<br>(50.4 – 64.7) | 17.6<br>(15.6 – 19.6) | 48.3<br>(42.7 – 53.9) | 12.1<br>(11.2 – 13.0) | 36.3<br>(31.7 – 41.0) |
| 14 – 17 years | Male | 51 | 12.9<br>(11.9 – 14.0) | 34.9<br>(32.2 – 37.6) | 17.4<br>(15.6 – 19.2) | 49.3<br>(44.0 – 54.6) | 15.1<br>(13.7 – 16.5) | 41.5<br>(37.6 – 45.5) | 10.4<br>(9.8 – 11.0) | 30.5<br>(28.2 – 32.7) |
|  | Female | 64 | 11.2<br>(10.4 – 12.1) | 31.8<br>(29.5 – 34.0) | 15.6<br>(14.0 – 17.3) | 43.9<br>(39.3 – 48.5) | 13.9<br>(12.6 – 15.2) | 37.9<br>(34.4 – 41.4) | 10.6<br>(9.9 – 11.3) | 31.9<br>(28.7 – 35.2) |

