## Supplementary material for "Reference values for amplitude-integrated EEGs in children from 1 month to 17 years of age": Pocket card

### Reference values for pediatric aEEG

##### Channels according to the 10-20 system

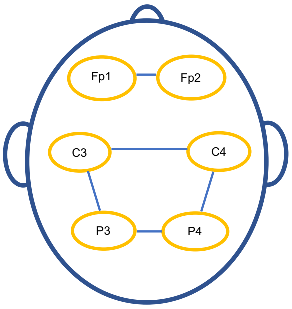


For best comparability, locate the aEEG electrodes according to one or several of the channels above for best comparability

##### Amplitude estimation

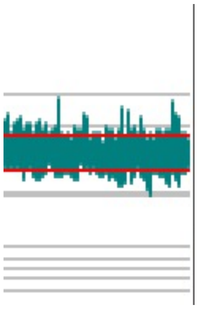

To assess the upper and lower amplitudes, try to estimate the margins of the main aEEGs, ignore solitary spikes and sections with artifacts

These percentiles were deduced from awake healthy children. Use in critically ill, asleep or sedated children requires cautious evaluation of comcomitant circumstances that may alter EEG activity or affect the plausibility of the measurement!

### Percentile values

| Age | Channel | | Percentile 10th | 25th | 50th | 75th | 90th |
|---|---|---|---|---|---|---|---|
| < 1 year | C3-P3/C4-P4 | Lower | 11 | 13 | **16** | 21 | 25 |
| | | Upper | 34 | 42 | **51** | 62 | 97 |
| | C3-C4 | Lower | 14 | 16 | **19** | 24 | 27 |
| | | Upper | 43 | 50 | **56** | 68 | 82 |
| | P3-P4 | Lower | 13 | 14 | **18** | 22 | 27 |
| | | Upper | 39 | 47 | **61** | 72 | 106 |
| | Fp1-Fp2 | Lower | 11 | 14 | **16** | 23 | 25 |
| | | Upper | 36 | 59 | **72** | 96 | 106 |
| 1 year | C3-P3/C4-P4 | Lower | 15 | 18 | **20** | 23 | 25 |
| | | Upper | 40 | 48 | **56** | 62 | 77 |
| | C3-C4 | Lower | 20 | 22 | **24** | 27 | 28 |
| | | Upper | 51 | 55 | **65** | 77 | 85 |
| | P3-P4 | Lower | 17 | 19 | **23** | 25 | 29 |
| | | Upper | 48 | 53 | **60** | 77 | 85 |
| | Fp1-Fp2 | Lower | 14 | 16 | **19** | 23 | 27 |
| | | Upper | 49 | 62 | **72** | 86 | 129 |
| 2 - 5 years | C3-P3/C4-P4 | Lower | 12 | 15 | **18** | 20 | 25 |
| | | Upper | 30 | 39 | **46** | 56 | 67 |
| | C3-C4 | Lower | 16 | 19 | **23** | 26 | 31 |
| | | Upper | 41 | 47 | **56** | 70 | 81 |
| | P3-P4 | Lower | 16 | 19 | **22** | 26 | 32 |
| | | Upper | 42 | 49 | **57** | 70 | 86 |
| | Fp1-Fp2 | Lower | 14 | 16 | **19** | 23 | 25 |
| | | Upper | 47 | 55 | **71** | 88 | 113 |
| 6 - 9 years | C3-P3/C4-P4 | Lower | 11 | 12 | **15** | 19 | 24 |
| | | Upper | 28 | 35 | **42** | 53 | 60 |
| | C3-C4 | Lower | 14 | 15 | **19** | 23 | 28 |
| | | Upper | 34 | 41 | **53** | 63 | 72 |
| | P3-P4 | Lower | 15 | 17 | **21** | 27 | 33 |
| | | Upper | 39 | 48 | **61** | 74 | 93 |
| | Fp1-Fp2 | Lower | 11 | 13 | **15** | 16 | 20 |
| | | Upper | 33 | 38 | **44** | 56 | 75 |
| 10 - 13 years | C3-P3/C4-P4 | Lower | 9 | 10 | **14** | 19 | 23 |
| | | Upper | 24 | 29 | **38** | 52 | 61 |
| | C3-C4 | Lower | 12 | 14 | **17** | 22 | 26 |
| | | Upper | 30 | 37 | **49** | 57 | 74 |
| | P3-P4 | Lower | 14 | 16 | **21** | 26 | 31 |
| | | Upper | 35 | 43 | **53** | 73 | 90 |
| | Fp1-Fp2 | Lower | 9 | 10 | **12** | 14 | 17 |
| | | Upper | 22 | 28 | **34** | 45 | 53 |
| 14 - 17 years | C3-P3/C4-P4 | Lower | 7 | 9 | **11** | 15 | 19 |
| | | Upper | 17 | 23 | **31** | 40 | 52 |
| | C3-C4 | Lower | 9 | 11 | **14** | 17 | 21 |
| | | Upper | 23 | 29 | **37** | 49 | 58 |
| | P3-P4 | Lower | 9 | 12 | **15** | 20 | 26 |
| | | Upper | 26 | 31 | **43** | 59 | 71 |
| | Fp1-Fp2 | Lower | 8 | 9 | **10** | 12 | 14 |
| | | Upper | 21 | 24 | **29** | 37 | 45 |

Pattern legend ·············· 10th — — — — 25th ——— 50th — — — — 75th ·············· 90th

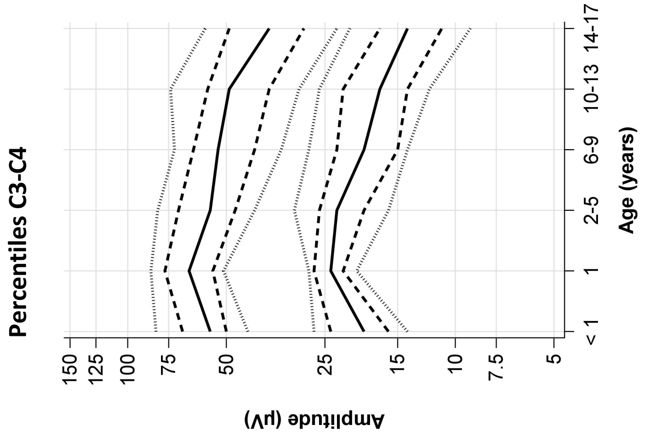


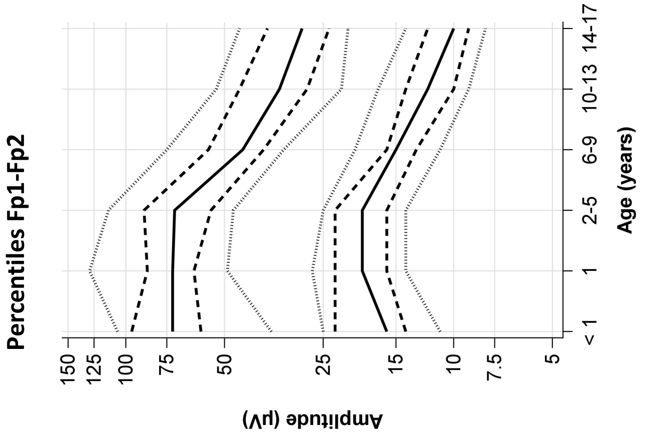


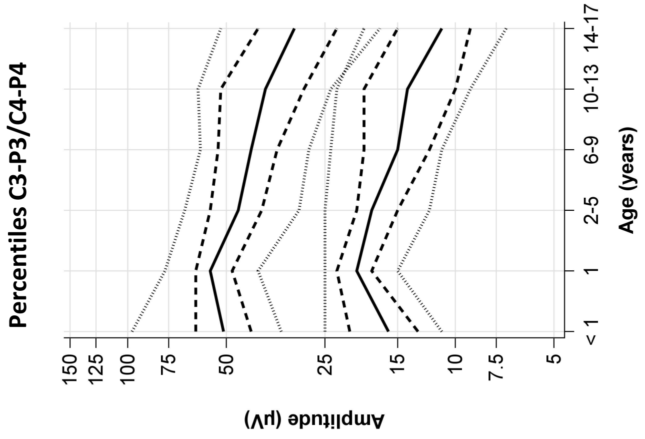


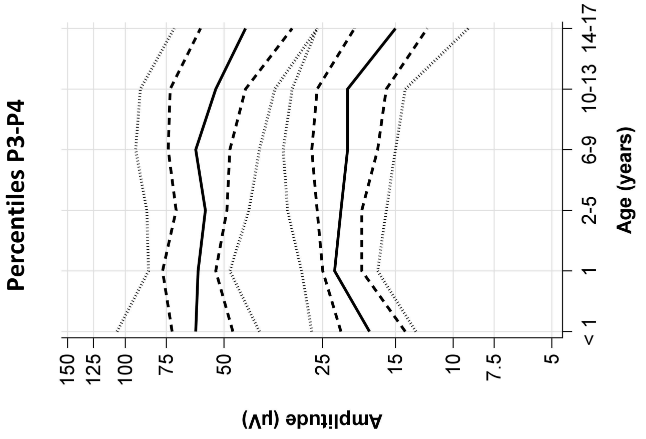
